# AI-Driven Smartphone Screening for Acute COPD Exacerbations: A Non-Self-Report Approach to Improve Health Equity in Developing Regions

**DOI:** 10.1101/2025.06.28.25330256

**Authors:** Yanbin Gong, Chi Xu, Caixia Mo, Jianhui Wu, Huiyong Lin, Huaan Su, Qingpeng Zhang, Qian Zhang, Shifang Yang

## Abstract

Acute exacerbations of chronic obstructive pulmonary disease (AECOPD) are critical clinical events that necessitate prompt intervention. However, their detection remains challenging in primary care settings, especially in resource-limited regions. A lack of disease awareness among both patients and healthcare providers often leads to delayed diagnoses and suboptimal management. To address this issue, we developed an AI-based AECOPD detection system that leverages standard mobile phone microphones for auscultation, specifically designed for novice users and eliminating the need for subjective symptom assessments or patient-reported scales. Our system demonstrated robust performance in automatically detecting exacerbations, achieving an area under the curve (AUC) of 0.955 (95% CI: 0.929-0.976). This research highlights the potential of AI-driven solutions to enhance COPD management in underserved populations with limited access to specialist medical resources, thereby promoting health equity. The digital health system shows promise for improved long-term management of COPD and is projected to save 41.81 billion CNY (Median: 24.25 billion, 95% CI: -7.75-198.33 billion), particularly benefiting primary care settings where access to pulmonary specialists is constrained.

## 1 Introduction

Chronic obstructive pulmonary disease (COPD) poses a significant global health challenge, ranking as a leading cause of morbidity and mortality worldwide [1]. In China alone, nearly 100 million individuals are affected by COPD, accounting for approximately one-quarter of the global patient population and imposing a substantial economic burden [2].

Chronic obstructive pulmonary disease (COPD) poses a significant global health challenge, ranking as a leading cause of morbidity and mortality worldwide [1]. In China alone, nearly 100 million individuals are affected by COPD, accounting for approximately one-quarter of the global patient population and imposing a substantial economic burden [2]. Acute exacerbations (AE) of COPD (AECOPD) are critical events that significantly increase mortality risk, accelerate lung function decline, diminish quality of life of COPD patients, and hospitalizations following AE account for over 70% of COPD-related medical costs [3, 4]. In China, patients with COPD experience on average 0.5-3.5 AEs annually, with treatment costs increasing from US$3,472 to US$4,538 between 2009 and 2017 in Beijing [5].

Effective management, including appropriate and timely treatment, is crucial to mitigate the risk and impact of AECOPD [6]. However, primary care services, the central to COPD ongoing monitoring, often lack sufficient knowledge about AE diagnosis and management [7]. Studies indicate that patients managed solely by primary care physicians frequently receive treatment that does not fully align with the Global Initiative for Chronic Obstructive Lung Disease (GOLD) recommendations [8, 9]. In China, knowledge of AECOPD among healthcare providers remains insufficient [10]. For instance, a 2015 survey in Shanghai, one of China’s most developed regions, revealed that only 19.4% of 593 general practitioners understood the distinction between AE and stable states of COPD [11]. The treatment given during different states deviate from international standards, which could partly explain the high AE rate in developing areas [12].

Patient-related factors further complicate these challenges. Many COPD patients, particularly those with lower educational levels in developing regions, often lack a thorough understanding of their condition and struggle to accurately report symptoms. This leads to poor self-management practices that can increase the risk of AE [13, 14]. While symptom assessment tools like EXACT-Pro exist [15], their effectiveness is limited by these patient factors and intra-individual variability [16, 17]. Additionally, a significant proportion of COPD patients (16–57%) experience some degree of cognitive impairment, which can hinder accurate self-reporting of their condition [18]. Besides, patients may under-report symptoms due to denial, embarrassment, or limited health literacy [19]. These patient-related factors, coupled with disparities in healthcare resource allocation, contribute to significantly higher AECOPD rates in rural areas and secondary hospitals compared to urban tertiary hospitals [20], a disparity also noted in broader healthcare delivery observations.

The emergence of telemedicine offers potential for enhancing health equity, particularly in developing areas [21–24]. However, remote management of COPD presents unique challenges. Unlike cardiovascular conditions where patients can report objective measurements, COPD monitoring relies heavily on subjective symptom reporting and availability of experienced physicians for remote assessment [25]. Recently, ResApp (now Pfizer Inc.) developed a smartphone-based algorithm for AECOPD diagnosis using cough audio data combined with patient-reported features such as age, fever, and new cough [26]. However, this system’s reliance on patient-reported symptoms potentially limits its utility in developing areas, and its sensitivity of 82.6% (95% CI: 72.9-89.9%) and specificity of 91.0% (95% CI: 82.4-96.3%) may generate an additional medical cost burden.

This study is motivated by the critical need for an accessible, objective, and cost-effective method to detect AECOPD, thereby advancing health equity in COPD management for underserved populations. As shown in Fig. 1(a), we developed a smartphone-based AI system that detects AECOPD by utilizing the device’s integrated microphone to capture and analyze lung sounds, eliminating reliance on patient-reported symptoms. Patients are guided to perform standardized deep breathing and coughing maneuvers, mirroring auscultation during clinical visits. While previous work demonstrated smartphone-based cardiac auscultation [27, 28], lung auscultation presents greater challenges due to lower signal-to-noise ratios [29]. Our system addresses this by integrating clinical physiological insights with a sophisticated AI model. The design is based on the understanding that AECOPD is characterized by alterations in physiological parameters such as dyspnea, respiratory rate, heart rate, oxygen saturation, and inflammatory markers [30]. Although direct measurement of all these parameters via smartphone audio is not feasible, their manifestations are often strongly correlated with discernible acoustic features [31, 32]. Deep-breathing auscultation provides greater diagnostic detail compared to cough analysis, yet it presents technical challenges due to inherently lower signal amplitude, potentially resulting in unfavorable signal-to-noise ratios (SNR). Crucially, recent advancements in AI-driven audio signal processing, particularly the foundation model OPERA, [33] pre-trained on hundreds of thousands of diverse audio samples, enable our system to extract meaningful patterns from low-SNR smartphone recordings.

**Fig. 1:**
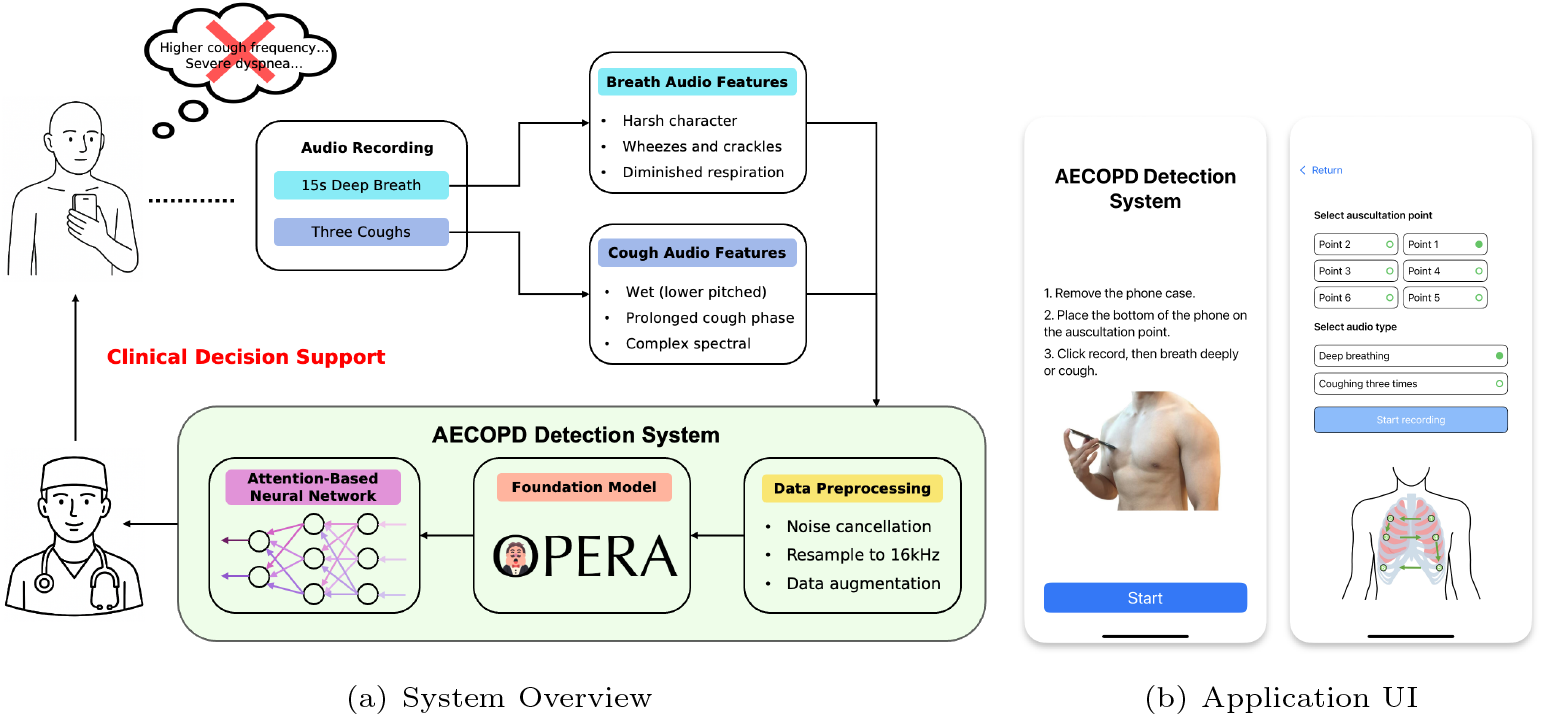
AI-System Overview and Usage Process. **(a)** Overall system architecture diagram. The process starts with recording the patient’s breathing and coughing audio, and then extracting key acoustic features from them. After preprocessing, these data are input into a neural network based on the attention mechanism and the OPERA basic model for analysis, ultimately providing doctors with clinical decision support for AECOPD treatment. **(b)** The user interface of the smartphone application for audio collection. The interface design is simple and the operation instructions are clear (such as selecting auscultation points, selecting audio types), demonstrating the ease of use of the tool.

**Fig. 2:**
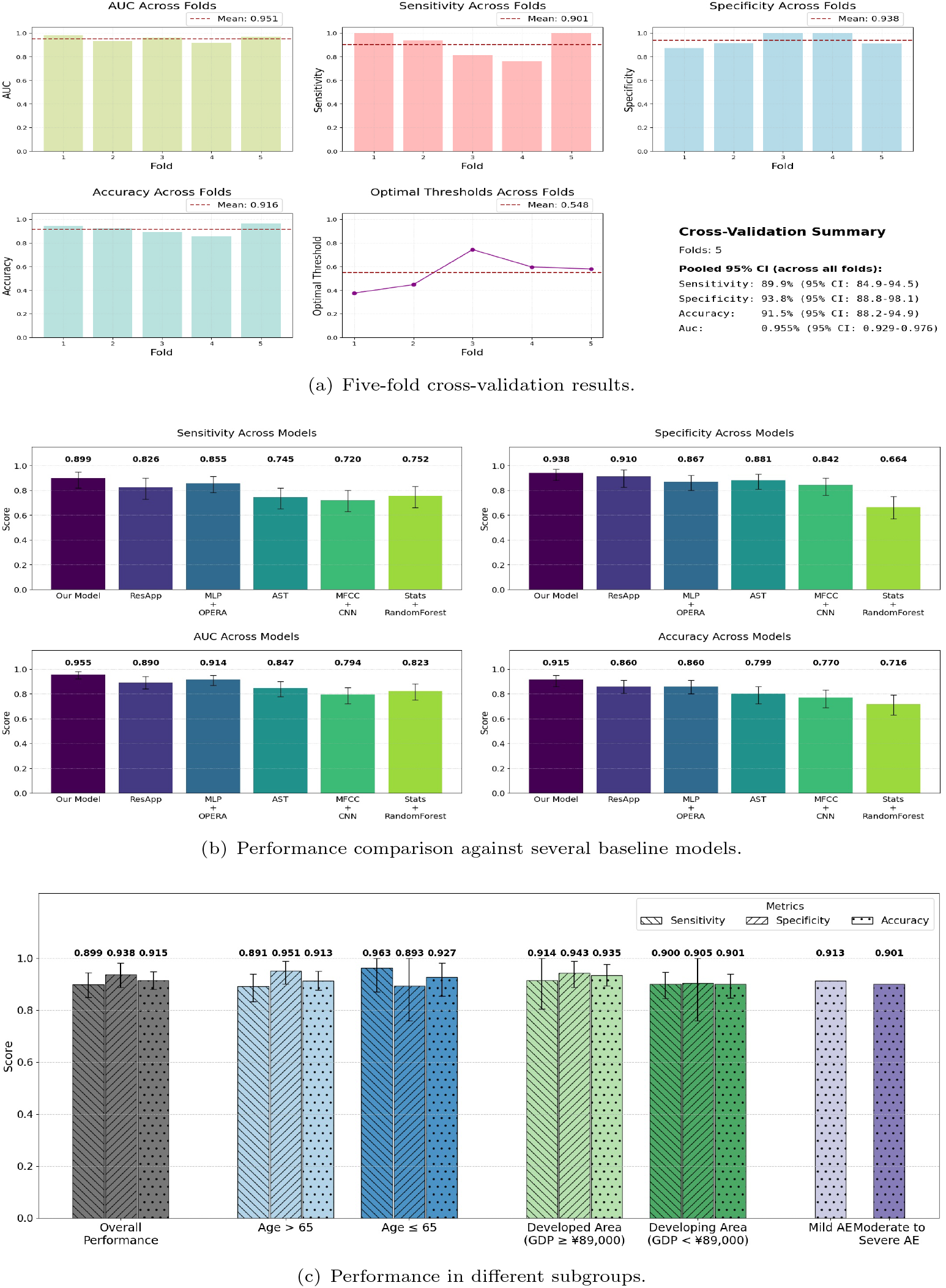
Overview of AI system performance. **(a)** Five-fold cross-validation results showing stable performance across key metrics. **(b)** Performance comparison against several baseline models across four different metrics (Sensitivity, Specificity, AUC, Accuracy), demonstrating the superiority of our proposed model. **(c)** Consistent performance across diverse demographic and socioeconomic subgroups supports more equitable healthcare delivery.

**Fig. 3:**
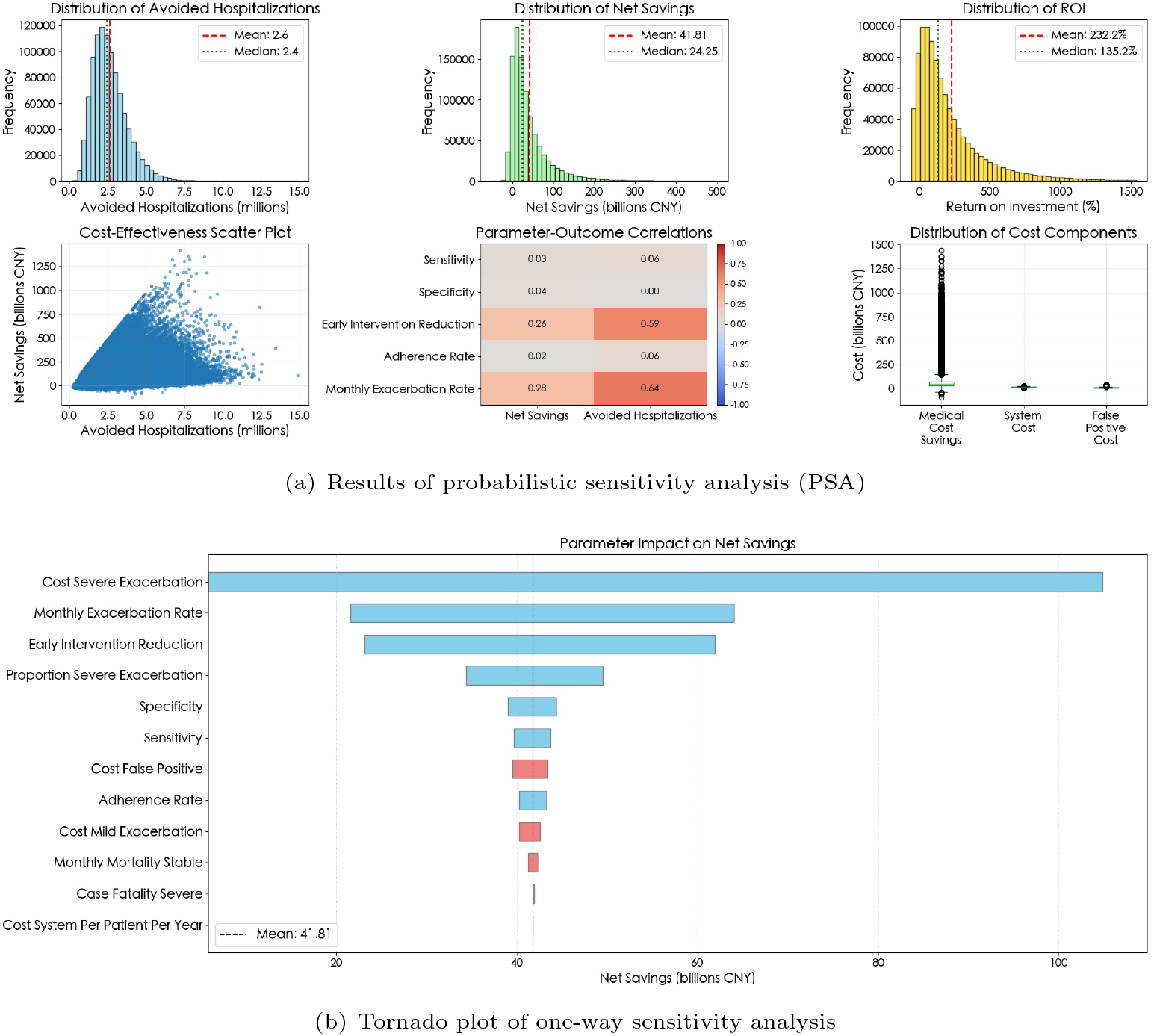
Health economics of AECOPD detection system. **(a)** Results of probabilistic sensitivity analysis (PSA). The probability distribution of the model’s economic output was right-skewed, predicting an average of 2.6 million hospitalizations avoided, a net saving of RMB 41.81 billion, and a return on investment (ROI) of 232.2%. Net savings were strongly positively correlated with the number of hospitalizations avoided. Parameter-output correlation analysis showed that early intervention effects and monthly acute exacerbation rates were key factors in determining the model’s output. **(b)** Tornado plot of the one-way sensitivity analysis. This plot reveals the sensitivity of net savings to key parameters, with severe exacerbation costs, monthly exacerbation rates, and early intervention effectiveness being the most important drivers of economic benefits.

Fig. 1(b) illustrates a typical use case of our mobile application. Users are guided by the mobile application to position the smartphone on specific chest locations to perform sequential deep breathing and coughing maneuvers, which is user friendly for elderly people. The system automatically processes recordings and allows re-recording if compromised by misplacement or external disturbances, making it user-friendly for individuals with limited education or cognitive impairments.

To develop and validate this system, we conducted a multi-center study involving one tertiary hospital and two community hospitals in Guangzhou, and eight county-level hospitals. User studies demonstrates the system’s ease of use for novice users. The system achieved an Area Under the Curve (AUC) of 0.955 (95% CI: 0.929–0.976) for AECOPD detection. We estimate that widespread adoption of this technology could yield substantial health economic benefits, projected at 41.81 billion CNY (Median: 24.25 billion, 95% CI: -7.75-198.33 billion), by improving health equity and alleviating the healthcare burden in developing regions.

## 2 Results

### 2.1 Study Population and Demographics

This prospective, multi-center study was conducted in 13 hospitals in central and southern China from November 2024 to June 2025. A total of 292 COPD patients were recruited at the beginning of the study. After excluding 18 patients with missing or damaged data, a total of 274 patients were finally included in the analysis cohort for model development and validation. The cohort included 165 patients with AECOPD and 109 patients in the stable state. The baseline demographics and clinical characteristics of the study population are detailed in Table 1. Statistically significant differences were observed between the AECOPD group and the stable group in terms of age, smoking status, and number of AE in the last year (all p *<*0.001). Specifically, patients in the AECOPD group were older, had a higher percentage of current smokers, and reported more AE in last year compared with the stable group. No significant difference was found in the gender distribution between the two groups.

**Table 1:** Baseline Demographics and Clinical Characteristics of the Study Population.

| Characteristic | All (274) | AECOPD (165) | Stable (109) | p-value |
| --- | --- | --- | --- | --- |
| Subjects, n | 274 | 165 | 109 |  |
| **Age (years)** |  |  |  | 0.001 |
| Mean ± SD | 72.9 ± 8.8 | 74.3 ± 8.5 | 70.8 ± 8.8 | |
| Median (Q1, Q3) | 74.0 (67.0, 79.0) | 75.0 (69.0, 81.0) | 71.0 (65.0, 78.0) |  |
| **Sex, n (%)** |  |  |  | 0.621 |
| Male | 221 (80.7%) | 131 (79.4%) | 90 (82.6%) |  |
| Female | 53 (19.3%) | 34 (20.6%) | 19 (17.4%) |  |
| **Smoking status, n (%)** |  |  |  | 0.000 |
| Current smoker | 78 (28.5%) | 72 (43.6%) | 6 (5.5%) |  |
| Former smoker | 192 (70.1%) | 90 (54.5%) | 102 (93.6%) |  |
| Never smoker | 4 (1.5%) | 3 (1.8%) | 1 (0.9%) |  |
| **Exacerbation Count** |  |  |  | 0.000 |
| Median (Q1, Q3) | 1.0 (0.0, 2.0) | 1.0 (1.0, 2.0) | 0.0 (0.0, 1.0) |  |

### 2.2 Dataset Composition

Data collection was designed to reflect real-world clinical practice. The process was integrated into routine auscultation of outpatients and performed at the bedside of inpatients in the ward, which are non-isolated clinical environments with inevitable ambient noise. Each patient’s condition was diagnosed by two independent medical professionals as stable COPD, mild AE, or moderate to severe AE (requiring hospitalization). During our clinical visits, 142 patients presented with moderate-to-severe AE, which reflects the trend of patients seeking treatment later in the course of AE in our region. Data were collected by researchers at the 2nd, 4th, and 6th intercostal spaces with an average collection time of 4 minutes. The algorithm was well tolerated, with 88.0% of patients describing it as simple and convenient, and only 5.1% considering it unsuitable for daily use.

### 2.3 AI system accurately classifies disease states using only audio data

To evaluate the performance of the system and ensure its robustness, this study used a five-fold stratified cross-validation strategy, with stratification based on the patient’s clinical status to ensure that each subset maintained the same proportion of patient group representation as the original dataset. As shown in Fig. **??**, the system demonstrated excellent discriminative power, with a pooled AUC of 0.955 (95% CI: 0.929–0.976) across all five-fold cross-validations. This performance was consistent across metrics, including 91.5% pooled accuracy (95% CI: 88.2–94.9%), 89.9% sensitivity (95% CI: 84.9–94.5%), and 93.8% specificity (95% CI: 88.8–98.1%). The consistency of the results across the data groups confirmed the stability of the model performance. Although the optimal decision thresholds varied slightly between groups (mean: 0.548 ± 0.128), the average AUC for each group was still as high as 0.951, with minimal differences in accuracy (0.916 ± 0.039) and F1 score (0.924 ± 0.039). Notably, the model achieved 100% specificity in two of the five groups, indicating that it can accurately identify patients with stable disease and minimize false positives.

To further validate our model, we comprehensively compare its performance with five baseline models covering different technical paths. As shown in Fig. **??**(b), our model significantly outperforms all the compared models on all key evaluation metrics. Specifically, we first compare our model with a variant that uses the same OPERA acoustic embedding features but uses a standard multi-layer perceptron (MLP) for classification. Our model achieves significant improvements in AUC (0.955 vs 0.914), sensitivity (0.899 vs 0.855), specificity (0.938 vs 0.867), and accuracy (0.915 vs 0.860). This result strongly demonstrates that the unique architectural design of our model can more effectively learn and extract disease-related discriminative features from OPERA embeddings. In addition, we also compare with the ResApp model, which not only analyzes cough audio but also combines patient self-report data. Despite ResApp’s exploitation of additional multimodal information, our audio-only model still demonstrates stronger performance in terms of AUC (0.955 vs 0.890), sensitivity (0.899 vs 0.826), and specificity (0.938 vs 0.910). This highlights the technical advantages and potential of our approach in making accurate diagnoses relying solely on audio signals. Finally, our model outperforms commonly used methods in the field of audio-based disease classification models, such as the fine-tuned Audio Spectrogram Transformer (AST) [34], using convolutional neural networks to process Mel-frequency cepstral coefficients (MFCC+CNN), or using random forests with acoustic statistical features. Our method performs much better than the best baseline model AST (AUC 0.847, accuracy 0.799), demonstrating its great potential as an innovative disease screening and monitoring tool.

### 2.4 Consistent and equitable performance in heterogeneous patient populations

To analyze the generalization ability of the model, a series of pre-specified stratified subgroup analyses were performed on the entire cohort of 274 patients based on key clinical and socioeconomic characteristics. The diagnostic accuracy of the model remained consistent across the range of AECOPD severity. In the cohort diagnosed with AECOPD, the system successfully identified 91.3% of mild AE (n=23) and 90.1% of moderate to severe AE (n=142). This consistently high level of performance, especially in mild cases that are difficult to diagnose, highlights its potential for early detection of the disease. The system showed robust performance across patients of different ages. In patients older than 65 years (n=219), the model had an accuracy of 91.3% (95% CI: 87.2%–94.5%), a sensitivity of 89.1% (95% CI: 83.2%–93.9%), and a specificity of 95.1% (95% CI: 89.8%–98.9%). In the younger cohort (n=55), performance was comparable, with accuracy, sensitivity, and specificity of 92.7% (95% CI: 85.5%–98.2%), 96.3% (95% CI: 87.5%–100.0%), and 89.3% (95% CI: 75.9%–100.0%), respectively. Analysis of the misclassified samples provided further insights false-negative samples in the older cohort occasionally occurred in patients with no significant abnormalities on clinical auscultation, suggesting that the model may have captured subtle subclinical signs of AE that are often masked by age-related physiological changes. In contrast, the few false-positive samples in the younger cohort were often associated with non-AE of respiratory inflammation, whose acoustic patterns may be similar to acute events. In addition, we also evaluated the application value of the system in different medical ecosystems by stratification by regional GDP per capita. The performance of the model showed excellent stability across these significantly different economic strata. In developed regions (GDP per capita *>*89,000 CNY ; n=123), the system’s accuracy was 93.5% (95% CI: 88.6%–97.6%). In developing regions (n=151), the system also achieved a comparable accuracy of 90.1% (95% CI: 84.8%–94.7%). The chi-square test analysis showed that the difference in accuracy between the two region groups did not reach a statistically significant level (*χ*^2^=1.037, P=0.309). This result indicates that the performance of the system is independent of regional economic factors and potential confounding variables, thus shows great potential in developing areas.

In summary, these subgroup analyses show that the model is not only accurate across disease severity, age, and socioeconomic status, but also highly robust, reliable, and fair. This provides strong evidence for its widespread and effective deployment in diverse real-world clinical settings.

### 2.5 Economic modeling indicates substantial cost reductions

To evaluate the health economic value of our system, we conducted a probabilistic sensitivity Monte Carlo analysis and compared it with a similar system, ResApp. The results showed that our model exhibited superior cost-effectiveness. On average, our system was expected to generate a net savings of 41.81 billion CNY (Median: 24.25 billion, 95% CI: -7.75-198.33), avoid 2.62 million hospitalizations (median 2.44 million, 95% CI: 981,311-5,326,055), and achieve a 135.18% return on investment (median). In contrast, although the comparator model also showed positive economic benefits, its indicators were lower than our system, and our system could bring 21.5% higher net savings. More importantly, our system performs more robustly under uncertainty, with a probability of achieving cost savings of 89.0%, significantly better than the 81.8% of the comparator model, which means that it has a higher certainty of achieving economic benefits in actual applications. One-way sensitivity analysis further revealed that the benefits of the model are mainly driven by key parameters such as severe AECOPD cost, early intervention cost reduction, and monthly AE rate, confirming its core mechanism of achieving cost control through effective intervention. And the drawback is from cost of false positive and cost difference between the outpatient visit and in-hospital. Overall, our system not only has a clear advantage in the expected value of economic benefits, but its robustness also ensures higher value potential in real-world applications.

## 3 Discussion

In this study, we have developed and validated a smartphone-based AI system that can objectively detect AECOPD using only deep breathing and coughing sounds captured by a microphone. Our five-fold cross-validation demonstrated excellent discrimination ability, with a pooled AUC of 0.955 (95% CI: 0.929–0.976), and achieved 91.5% accuracy (95% CI: 88.2–94.9%), 89.9% sensitivity (95% CI: 84.9–94.5%), and 93.8% specificity (95% CI: 88.8–98.1%). Notably, the system exhibited robust and fair performance across diverse patient subgroups, including variations in disease severity, age, and socioeconomic status, underscoring its strong generalization capability.

A key advantage of our system is its capacity to advance health equity through its inherent usability and objectivity. By leveraging the ubiquitous platform of smartphones, it provides an accessible and easily operable tool for AECOPD detection, directly bridging the urban-rural healthcare divide common in low-resource settings. This approach overcomes the reliance on specialized patient and health provider knowledge in traditional assessments, while its operational simplicity ensures accessibility for individuals with limited education or cognitive impairments. The economic implications are also great. Our system is estimated to generate net savings of 41.81 billion CNY (5.846 billion USD) in China, which can substantially alleviate the national healthcare burden and reduce costs for the underserved populations. Collectively, these findings position our system as a robust, scalable tool for widespread deployment in primary care, presenting a pathway to achieving health equity in COPD management.

Moreover, this work signifies a transition from subjective assessments to objective remote chronic disease management. Our methodology decouples clinical monitoring from the uncertain subjective interpretations of patients, establishing a high-fidelity objective data record. This analytical framework can be fully extended to other disease monitoring using sound signals as biomarkers, such as the wheezing of asthma patients, the nighttime cough of heart failure patients, and even the changes in speech characteristics caused by neurodegenerative diseases. A more transformative potential lies in integrating our acoustic analysis into a patient-centered multimodal monitoring ecosystem. An attractive path is to integrate our respiratory sound analysis with other physiological assessment technologies that are also based on smartphones, such as heart sound auscultation technology based on mobile phone microphones [35], so as to achieve a synchronous and comprehensive assessment of the cardiopulmonary system. Additionally, incorporating data from wearable devices—such as activity levels, sleep patterns, and blood oxygen saturation—along with electronic logs of medication and symptoms, can create a detailed “digital phenotype” of a patient’s health trajectory. This low-cost, objective patient management method will benefit patients in areas with scarce medical resources.

To achieve the goal of detecting AECOPD using only audio data collected by smartphones, the core design of this system is to combine advanced artificial intelligence technology with deep clinical physiological insights. Our design is based on the clinical physiological principle that AECOPD is characterized by changes in a series of physiological parameters such as dyspnea, respiratory rate, and inflammatory markers. Although smartphones cannot directly capture these indicators, the respiratory acoustic features caused by these physiological changes, such as wheezes and diminished breath sounds, are closely related to them, which provides a theoretical basis for audio detection. However, direct lung auscultation using smartphones faces the challenge of limited available data with low SNR. To overcome this problem, we leverage the power of advanced audio foundation model OPERA, which is pre-trained on hundreds of thousands of diverse audio samples, enabling it to accurately extract meaningful patterns related to AECOPD from low SNR ratio recordings, effectively solving the bottleneck of data quality and quantity. The designated pre and post foundation model processing also play a vital role in helping the system gets better result. In addition, the system design has good scalability. With the development of smartphone technology, it can be easily connected to external devices such as digital stethoscopes in the future to obtain higher quality audio data, further improving the performance and application potential of the system.

Despite these promising results, there are limitations to our study.

The “gold standard” for diagnosing AECOPD is not entirely objective and relies on clinician evaluations, which inherently include subjective judgments. This implies that some patterns learned by our system may reflect clinician diagnostic preferences rather than purely pathophysiological states, suggesting that the model’s performance ceiling is constrained by existing clinical diagnostic criteria. Additionally, our study population primarily consisted of individuals from central and southern China, which may limit the generalizability of our findings to other COPD patient groups. Furthermore, we observed a notable disconnect between patients’ self-reported histories of AE and their objective disease severity. Due to the widespread interoperability barriers of electronic medical record systems, the acquisition of AE history is forced to rely on patients’ subjective recollections. Many patients with severely impaired lung function reported an abnormally low frequency of AE, likely due to their inability to recognize or define these events. When prompted by specific medical resource utilization (such as hospitalization or intravenous treatment), many patients who initially denied the existence of AE events were able to confirm the existence of AE events. Therefore, patient recollection is not only an unreliable proxy, but is also likely to lead to systematic misestimation of the true activity of the disease. In addition, the performance differences of different models of smartphone microphones and the potential impact of environmental noise are common issues that need to be considered when collecting medical data based on consumer-grade devices. Finally, this study lacks longitudinal monitoring data to evaluate the performance of the system in long-term use, and the health economic analysis is based on model assumptions, and its benefits in real-world practice still need to be verified by prospective studies.

To further validate and expand the clinical value of this system, future research should focus on the following key directions. First, we plan to conduct a large-scale, prospective, real-world validation study to allow patients to use the system in their home environment and for health providers to deploy it in primary care institutions to evaluate its actual effectiveness in an uncontrolled environment. Second, the system’s potential for application in longitudinal monitoring of COPD disease status and early prediction of AE should be explored, which may provide a new way to achieve preemptive intervention. Third, integrating the system with electronic health records (EHR) and digital pulmonary rehabilitation program is a big step in achieving datadriven personalized chronic disease management. We also plan to directly evaluate the actual impact of the system on clinical decision-making and patient outcomes through interventional clinical trials, and conduct more precise health economic evaluations based on real-world usage data.

In summary, this study successfully developed and validated an objective, convenient, and highly accessible smartphone-based AI screening for AECOPD. The core contribution of this work is that it provides an objective decision support tool for COPD patients and primary care community in resource-limited areas, thereby empowering health equity. This system greatly promotes the implementation of telemedicine strategies for COPD management. By achieving earlier detection of AECOPD and timely initiation of symptomatic treatment, it is expected to significantly reduce the severity of the disease, shorten the course of the disease, and reduce hospitalization rates. These clinical benefits will ultimately be transformed into effective relief of the socioeconomic burden associated with COPD through more efficient management of AE. Therefore, this system not only provides strong technical support for promoting health equity in the field of respiratory diseases, but also demonstrates a feasible path to transform the traditional chronic disease management system using mobile health technology, and is expected to improve the quality and accessibility of care for COPD patients worldwide.

## 4 Methods

### 4.1 Study Design and Setting

We conducted a prospective, multicenter cross-sectional study in central and southern China to develop and validate the AECOPD detection system. The study was conducted in multiple medical centers, including Guangdong Provincial People’s Hospital. All participants were fully informed of the purpose, procedures, potential risks and benefits of the study before enrollment and voluntarily signed written informed consent. The subjects of the study were recruited from the respiratory outpatient clinic and inpatient department of each participating center. The recruitment process was arranged during the outpatient consultation and clinical assessment. The data collected included auscultation sounds, basic information of the patients, and a COPD management questionnaire based on the subjective completion of the patients (see Appendix). The inclusion criteria were: 1) aged between 18 and 90 years; 2) confirmed diagnosis of COPD according to the diagnostic criteria of GOLD [36]; 3) able to cooperate with the completion of pulmonary function tests and related questionnaires; 4) able to cooperate with the use of inhaled medications; 5) not participating in other pulmonary rehabilitation programs within six months before enrollment. Exclusion criteria include patients with other severe respiratory diseases (such as severe pneumothorax, pulmonary embolism, etc.), uncontrolled severe systemic diseases (such as unstable cardiovascular disease, untreated tumors, etc.), musculoskeletal or nervous system diseases that affect movement, mental or cognitive disorders, and patients with a history of lung surgery within the past three months. This strict screening criteria is designed to ensure the safety of the research experiment and the homogeneity of the population.

### 4.2 Ethics statement

This study was conducted in compliance with the Declaration of Helsinki and received ethical approval from the Ethics Review Committee of Guangdong Provincial People’s Hospital (IRB no. KY2025-422-01). Written informed consent was obtained from all participants. An English translation of the ethics approval documentation is available from the corresponding author upon reasonable request.

### 4.3 Clinical Reference Standard

To ensure the accuracy and reliability of diagnostic labels, the clinical reference standard was established through a consensus agreement between two experienced respiratory physicians who independently assessed each participant. The clinical evaluations were performed in accordance with the 2021 Rome Proposal [30]. During the evaluation process, experts first distinguished whether the COPD patient was in a stable state or an AE state by whether the patient had dyspnea or worsening cough and sputum within 14 days, which may be accompanied by increased respiratory rate or heart rate. Subsequently, AECOPD patients were further divided into mild or moderate to severe based on a combination of objective physiological indicators, including dyspnea scores, vital signs, oxygen saturation, and C-reactive protein. Conflicts in evaluation opinions were discussed and negotiated to reach a final consensus. This process provides a definitive label for each sample as a benchmark for model development and validation.

### 4.4 Data Collection

A custom iOS application was developed for data collection in this study. The application can directly call the microphone at the bottom of the smartphone to collect the raw audio stream, thereby bypassing the interference caused by system-level signal processing optimized for voice calls (such as noise reduction and gain). During data collection, the subject remained seated, and the collector placed the bottom edge of the phone directly on the patient’s skin surface and collected respiratory sounds at symmetrical positions in the second, fourth, and sixth intercostal spaces. At each collection point, we recorded 15 seconds of deep breathing sounds and 3 spontaneous cough sounds in sequence.

### 4.5 Model Development

To extract discriminative features from complex acoustic signals, we implemented a series of processing steps and model designs. Initially, we applied Wiener filtering and bandpass filtering to preprocess the audio data, effectively removing both device noise and extraneous sounds from the breathing segments. Subsequently, we downsampled the data to 16 kHz and utilized the OPERA model [33] to encode the processed cough and deep breathing sounds into high-dimensional feature embeddings. Specifically, we employed two distinct encoders: OPERA-CT (768 dimensions), based on a Transformer architecture, and OPERA-CE (1280 dimensions), based on convolutional neural networks. This approach allowed us to convert each audio clip into a structured feature vector for subsequent modeling.

To integrate acoustic information from cough and breathing, we designed and implemented a multimodal fusion attention network. The model first maps CT and CE features of different dimensions to a unified feature space through independent projection layers. Then, the spatial dependency of different lung positions is captured by the intra-modal self-attention mechanism [37], and the cross-modal attention mechanism is used to achieve deep information interaction between cough and deep breathing features. Finally, the fused features are fed into a classifier to output the diagnosis probability of AECOPD.

To ensure the objectivity and robustness of the model evaluation, we adopted a 5-fold stratified cross-validation strategy based on “subject-visit” to divide the data set to strictly prevent data leakage. During the training phase, we only applied data enhancement techniques including Gaussian noise, feature inactivation and Mixup to the training set. The AdamW optimizer was used for model training, combined with cosine annealing learning rate scheduling. Considering the possible imbalance of sample categories, we selected Focal Loss as the loss function and used the Early Stopping mechanism to avoid overfitting.

In the model inference stage, we did not utilize a fixed classification threshold of 0.5. Instead, we determined the optimal probability threshold for each fold by maximizing Youden’s J statistic on its respective validation set, based on the analysis of the receiver operating characteristic (ROC) curve. This data-driven threshold provides a decisionmaking basis for subsequent performance evaluation (such as sensitivity, specificity, accuracy, etc.) and ensures that the model achieves a better balance between sensitivity and specificity.

### 4.6 Health Economic Analysis

We constructed a health economics analysis framework based on the state-transition Markov model to evaluate the potential economic value of the AECOPD detection system. The model includes four core health states: stable, mild AE, moderate-to-severe AE, and death. We simulated the natural course of the disease and clinical outcomes of the Chinese COPD patient population in a monthly cycle over a one-year period.

The model compared two scenarios: “standard of care” and “system-assisted management”. The key parameters of the model cover multiple dimensions such as epidemiology (such as the total population of COPD, the annual incidence of AE, and background mortality), clinical effectiveness (such as the effectiveness of early intervention in reducing severe AE), system effectiveness (such as the sensitivity and specificity of the system, and patient compliance) and economic costs (such as the cost of treating AE outpatient and inhospital, the cost of using the system, and the cost of false positive events). The baseline values and uncertainty ranges of these parameters are mainly derived from published literature, clinical expert consensus, and real-world data. The cost of treating inhospital AE is based on an empirical distribution constructed based on real hospitalization data from our center to enhance the model’s real-world fit.

We conducted a PSA to fully evaluate the impact of parameter uncertainty. In the analysis, all key parameters were assigned specific probability distributions, and 1 million simulations were performed using the Monte Carlo method. The main output indicators of the model include: net cost savings, avoided hospitalizations, avoided deaths, and ROI. In addition, we performed a one-way sensitivity analysis and presented it in the form of a tornado diagram to identify the key drivers that have the greatest impact on the model results.

### 4.7 Statistical Analysis

The training, validation, and testing of the models were performed with PyTorch and Scikit-learn. The performance metrics—including AUC, sensitivity, specificity, accuracy, and corresponding 95% confidence intervals were calculated with SciPy and NumPy. Figures were generated with Matplotlib and Seaborn.

## Supporting information

Implementation

## 4.8 Data availability

The datasets generated and analysed during the current study are not publicly available due to the privacy issue but are available from the corresponding author on reasonable request after anonymization.

### 4.9 Code availability

The underlying code for this study is not publicly available but may be made available to qualified researchers on reasonable request from the corresponding author.

## 5 Acknowledgments

The authors would like to thank the Hong Kong RGC for their financial support.

## 6 Author contributions

Q.Z., Q.Z. and S.Y. conceived and administered the study. Y.G. Y.G. performed the mobile application development, model development, baseline implementation, data analysis and curation. C.X. helped part of the model development. Y.G., Q.Z. and S.Y. analyzed the results. S.Y., C.M., J.W., H.L., H.S. and Q.Z. provided important clinical and social knowledge content for the study design. S.Y., C.M., J.W., H.L., H.S. and Y.G. conducted the clinical communication with patients and collected data. S.Y., C.M., J.W., H.L. and H.S. labelled data through a consensus agreement. Y.G. and S.Y. wrote the manuscript. Y.G., Q.Z., Q.Z., S.Y. and C.X. have critically revised the manuscript. All authors have critically read and approved the manuscript.

## 7 Competing interests

Qingpeng Zhang is an associate editor of npj Digital Medicine.

## Notes

### Competing Interest Statement

The authors have declared no competing interest.

### Funding Statement

This study was funded by Hong Kong RGC grant HB016, and 16206122.

### Author Declarations

Ehics committee of Guangdong Provincial Peoples Hospital gave ethical approval for this work(IRB no. KY2025-422-01)

## References

[1] Momtazmanesh, S., Moghaddam, S.S., Ghamari, S.-H., Rad, E.M., Rezaei, N., Shobeiri, P., Aali, A., Abbasi-Kangevari, M., Abbasi-Kangevari, Z., Abdelmasseh, M., et al.: Global burden of chronic respiratory diseases and risk factors, 1990–2019: an update from the global burden of disease study 2019. EClinicalMedicine 59 (2023)

[2] Wang, C., Xu, J., Yang, L., Xu, Y., Zhang, X., Bai, C., Kang, J., Ran, P., Shen, H., Wen, F., et al.: Prevalence and risk factors of chronic obstructive pulmonary disease in china (the china pulmonary health [cph] study): a national cross-sectional study. The Lancet 391(10131), 1706–1717 (2018)

[3] Anzueto, A.: Impact of exacerbations on copd. European Respiratory Review 19(116), 113–118 (2010)

[4] Mathioudakis, A.G., Janssens, W., Sivapalan, P., Singanayagam, A., Dransfield, M.T., Jensen, J.-U.S., Vestbo, J.: Acute exacerbations of chronic obstructive pulmonary disease: in search of diagnostic biomarkers and treatable traits. Thorax 75(6), 520–527 (2020)

[5] Liang, L., Shang, Y., Xie, W., Shi, J., Tong, Z., Jalali, M.S.: Trends in hospitalization expenditures for acute exacerbations of copd in beijing from 2009 to 2017. International journal of chronic obstructive pulmonary disease, 1165–1175 (2020)

[6] Wedzicha, J.A., Seemungal, T.A.: Copd exacerbations: defining their cause and prevention. The lancet 370(9589), 786–796 (2007)

[7] Yawn, B.P., Mintz, M.L., Doherty, D.E.: Gold in practice: chronic obstructive pulmonary disease treatment and management in the primary care setting. International journal of chronic obstructive pulmonary disease, 289–299 (2021)

[8] Vachon, B., Giasson, G., Gaboury, I., Gaid, D., Noël De Tilly, V., Houle, L., Bourbeau, J., Pomey, M.-P.: Challenges and strategies for improving copd primary care services in quebec: results of the experience of the compas+ quality improvement collaborative. International journal of chronic obstructive pulmonary disease, 259–272 (2022)

[9] Criner, R.N., Han, M.K.: Copd care in the 21st century: a public health priority. Respiratory care 63(5), 591–600 (2018)

[10] Sun, Y.-C.: Chronic obstructive pulmonary disease in primary healthcare institutions in China: Challenges and solutions. Chinese Medical Journals Publishing House Co., Ltd. 42 Dongsi Xidajie … (2020)

[11] Li, F., Cai, Y., Zhu, Y., Chen, X., Xu, X., Zhang, X., Yin, W., Zhu, W., Fu, H., Shen, C., et al.: The evaluation of general practitioners’ awareness/knowledge and adherence to the gold guidelines in a shanghai suburb. Asia Pacific Journal of Public Health 27(2), 2067–2078 (2015)

[12] Ko, F.W., Chan, K.P., Hui, D.S., Goddard, J.R., Shaw, J.G., Reid, D.W., Yang, I.A.: Acute exacerbation of copd. Respirology 21(7), 1152–1165 (2016)

[13] Wilkinson, T.M., Donaldson, G.C., Hurst, J.R., Seemungal, T.A., Wedzicha, J.A.: Early therapy improves outcomes of exacerbations of chronic obstructive pulmonary disease. American journal of respiratory and critical care medicine 169(12), 1298–1303 (2004)

[14] Wu, P., Jiang, Y.-q., Si, F.-l., Wang, H.-y., Song, X.-b., Sheng, C.-f., Xu, X., Li, F., Zhang, J.: Pharmaceutical treatment status of patients with copd in the community based on medical internet of things: a real-world study. NPJ Primary Care Respiratory Medicine 34(1), 10 (2024)

[15] Leidy, N.K., Wilcox, T.K., Jones, P.W., Murray, L., Winnette, R., Howard, K., Petrillo, J., Powers, J., Sethi, S., Group, E.-P.S., et al.: Development of the exacerbations of chronic obstructive pulmonary disease tool (exact): a patient-reported outcome (pro) measure. Value in health 13(8), 965–975 (2010)

[16] Leidy, N.K., Wilcox, T.K., Jones, P.W., Roberts, L., Powers, J.H., Sethi, S.: Standardizing measurement of chronic obstructive pulmonary disease exacerbations: reliability and validity of a patient-reported diary. American journal of respiratory and critical care medicine 183(3), 323–329 (2011)

[17] Alqahtani, J.S., Aquilina, J., Bafadhel, M., Bolton, C.E., Burgoyne, T., Holmes, S., King, J., Loots, J., McCarthy, J., Quint, J.K., et al.: Research priorities for exacerbations of copd. The Lancet Respiratory Medicine 9(8), 824–826 (2021)

[18] Chen, X., Yu, Z., Liu, Y., Zhao, Y., Li, S., Wang, L.: Chronic obstructive pulmonary disease as a risk factor for cognitive impairment: a systematic review and meta-analysis. BMJ Open Respiratory Research 11(1) (2024)

[19] Stellefson, M., Wang, M.Q., Campbell, O.: Factors influencing patient-provider communication about subjective cognitive decline in people with copd: Insights from a national survey. Chronic Respiratory Disease 21, 14799731241268338 (2024)

[20] Yang, T., Cai, B., Cao, B., Kang, J., Wen, F., Chen, Y., Jian, W., Wang, C.: Exacerbation in patients with stable copd in china: analysis of a prospective, 52-week, nationwide, observational cohort study (real). Therapeutic advances in respiratory disease 17, 17534666231167353 (2023)

[21] Liu, F., Jiang, Y., Xu, G., Ding, Z.: Effectiveness of telemedicine intervention for chronic obstructive pulmonary disease in china: a systematic review and meta-analysis. Telemedicine and e-Health 26(9), 1075–1092 (2020)

[22] North, M., Bourne, S., Green, B., Chauhan, A.J., Brown, T., Winter, J., Jones, T., Neville, D., Blythin, A., Watson, A., et al.: A randomised controlled feasibility trial of e-health application supported care vs usual care after exacerbation of copd: the rescue trial. NPJ digital medicine 3(1), 145 (2020)

[23] Liu, X., Chen, W., Qiu, Y., Li, X., Liu, F., Jiang, Z., Jia, F., Wang, C., Ji, R., Nawaz, T.R., et al.: Improving access to cardiovascular care for 1.4 billion people in china using telehealth. npj Digital Medicine 7(1), 376 (2024)

[24] Enichen, E.J., Heydari, K., Li, B., Kvedar, J.C.: Telemedicine expands cardiovascular care in china–lessons for health equity in the united states. npj Digital Medicine 8(1), 71 (2025)

[25] Velardo, C., Shah, S.A., Gibson, O., Clifford, G., Heneghan, C., Rutter, H., Farmer, A., Tarassenko, L., Heritage, E.C.T.A.T.V.W.M.H.C.T.L.J.S.R.J.P.L.: Digital health system for personalised copd long-term management. BMC medical informatics and decision making 17, 1–13 (2017)

[26] Claxton, S., Porter, P., Brisbane, J., Bear, N., Wood, J., Peltonen, V., Della, P., Smith, C., Abeyratne, U.: Identifying acute exacerbations of chronic obstructive pulmonary disease using patient-reported symptoms and cough feature analysis. npj Digital Medicine 4(1), 107 (2021)

[27] Kang, S.-H., Joe, B., Yoon, Y., Cho, G.-Y., Shin, I., Suh, J.-W., et al.: Cardiac auscultation using smartphones: pilot study. JMIR mHealth and uHealth 6(2), 8946 (2018)

[28] Luo, H., Lamata, P., Bazin, S., Bautista, T., Barclay, N., Shahmohammadi, M., Lubrecht, J.M., Delhaas, T., Prinzen, F.W.: Smartphone as an electronic stethoscope: factors influencing heart sound quality. European Heart Journal-Digital Health 3(3), 473–480 (2022)

[29] Williams, L., et al.: Auscultation Skills: Breath & Heart Sounds. Lippincott Williams & Wilkins, ??? (2009)

[30] Celli, B.R., Fabbri, L.M., Aaron, S.D., Agusti, A., Brook, R., Criner, G.J., Franssen, F.M., Humbert, M., Hurst, J.R., O’Donnell, D., et al.: An updated definition and severity classification of chronic obstructive pulmonary disease exacerbations: the rome proposal. American journal of respiratory and critical care medicine 204(11), 1251–1258 (2021)

[31] O’donnell, D., Parker, C.: Copd exacerbations· 3: pathophysiology. Thorax 61(4), 354–361 (2006)

[32] Melbye, H., Solis, J.C.A., Jácome, C., Pasterkamp, H.: Inspiratory crackles—early and late—revisited: identifying copd by crackle characteristics. BMJ Open Respiratory Research 8(1), 000852 (2021)

[33] Zhang, Y., Xia, T., Han, J., Wu, Y., Rizos, G., Liu, Y., Mosuily, M., Ch, J., Mascolo, C.: Towards open respiratory acoustic foundation models: Pretraining and benchmarking. Advances in Neural Information Processing Systems 37, 27024–27055 (2024)

[34] Gong, Y., Chung, Y.-A., Glass, J.: Ast: Audio spectrogram transformer. arXiv preprint arXiv:2104.01778 (2021)

[35] Ma, S., Chen, J., Ho, J.W.: An edge-device-compatible algorithm for valvular heart diseases screening using phonocardiogram signals with a lightweight convolutional neural network and self-supervised learning. Computer Methods and Programs in Biomedicine 243, 107906 (2024)

[36] Venkatesan, P.: Gold copd report: 2024 update. The Lancet Respiratory Medicine 12(1), 15–16 (2024)

[37] Vaswani, A., Shazeer, N., Parmar, N., Uszkoreit, J., Jones, L., Gomez, A.N., Kaiser, L., Polosukhin, I.: Attention is all you need. Advances in neural information processing systems 30 (2017)

