## Supplementary material for "AI-Driven Smartphone Screening for Acute COPD Exacerbations: A Non-Self-Report Approach to Improve Health Equity in Developing Regions": Implementation

June 28, 2025

#### 1 Model Development and Training Protocol

##### 1.1 Data Augmentation Strategy

To mitigate the risk of overfitting on a limited dataset and to enhance model robustness, we implemented a feature-level data augmentation strategy. Since the weights of the OPERA foundation models (OPERA-CT and OPERA-CE) were frozen during training, augmenting the resultant high-dimensional feature embeddings is a computationally efficient and direct approach to simulate variations in acoustic signals. This method exposes the downstream fusion network to a wider variety of data, encouraging it to learn more generalizable and invariant representations. For each sample in the training set, we dynamically generated augmented versions using a combination of the following techniques, as implemented in our Python script:

- **Gaussian Noise:** We introduced additive Gaussian noise with a mean of 0 and a randomly sampled standard deviation (e.g., between 0.01 and 0.05) to the feature vectors. This simulates the effect of background environmental noise and electronic noise from the recording device.
- **Dropout:** A random subset of feature dimensions was set to zero. This technique mimics the partial loss of acoustic signal and forces the model to not rely excessively on a small portion of discriminative features, thereby improving its robustness.
- **Feature Scaling:** The entire feature vector was multiplied by a random scalar drawn from a uniform distribution close to 1. This simulates variations in signal amplitude that may arise from differences in microphone proximity or the intensity of respiratory events.
- **Mixup:** We employed a simplified variant of Mixup by creating a linear interpolation between the original features and a random noise vector: $x' = \alpha x + (1 - \alpha)n$, where $x$ is the original feature vector, $n$ is a noise vector, and $\alpha$ is a coefficient sampled from a distribution close to 1. This helps to smooth the decision boundary and improve generalization.

##### 1.2 Model Architecture

Our classification model is a multimodal fusion attention network designed to integrate acoustic information from cough and deep breathing sounds. The architecture is composed of several key modules:

**Projection Layers.** Input features from OPERA-CT (768 dimensions) and OPERA-CE (1280 dimensions) are first mapped to a unified 256-dimensional space using two independent projection modules. Each module consists of a linear layer, followed by a `BatchNorm1d` layer for normalization, a `ReLU` activation function, and a `Dropout` layer with a rate of 0.5. This step is crucial for dimensional homogeneity and for stabilizing the training process.

**Intra-modal Self-Attention.** To capture the spatial dependencies among the six lung recording positions within each modality, we employ a multi-head self-attention mechanism (`nn.MultiheadAttention`). This module, configured with an embedding dimension of 256 and 4 attention heads, allows the model to weigh the importance of different lung locations and learn their relational patterns.

**Cross-modal Attention.** The core of our fusion strategy is a cross-modal attention mechanism, which facilitates deep information exchange between the cough and deep breathing modalities. We implement this with two parallel `nn.MultiheadAttention` modules. First, cough features serve as the query while deep breathing features act as the key and value, and vice versa in the second. This reciprocal attention allows each modality to be enriched by contextual information from the other.

**Aggregation and Fusion.** Following the attention stages, features from the six locations are aggregated into a single global feature vector for each modality. We utilize both global average pooling and global max pooling, summing their outputs to capture both the overall signal characteristics and the most salient features. The resulting 256-dimensional global vectors for CT and CE are then concatenated and passed through a final fusion layer, a linear layer from 512 to 256 dimensions with `BatchNorm1d`, `ReLU`, and `Dropout`, to produce a unified representation.

**Classifier.** The final 256-dimensional fused feature vector is fed into a two-layer feed-forward classifier. This classifier consists of a linear layer projecting from 256 to 128 dimensions, followed by `BatchNorm1d`, `ReLU`, `Dropout`, and a final linear layer that outputs logits for the two classes AE and stable. All linear layers in the network were initialized using the Xavier uniform distribution.

#### 1.3 Training and Evaluation Algorithm

The complete training and evaluation pipeline, incorporating the architecture and protocols described, is summarized in Algorithm 1. This process details the K-fold cross-validation loop, model training, optimal threshold determination, and final performance aggregation.

#### 1.4 Model Training and Optimization

Model training was conducted using the PyTorch framework, with all computations accelerated on a CUDA-enabled GPU. A fixed random seed (42) was used across all libraries (PyTorch, NumPy, random) to ensure the reproducibility of our experiments. The key hyperparameters and settings for our training protocol are summarized in Table 1.

#### 1.5 Evaluation Protocol

To ensure an unbiased and robust assessment of our model's performance, we adhered to a rigorous evaluation protocol.

**Cross-Validation Strategy.** A 5-fold stratified cross-validation protocol was employed. Critically, stratification was performed at the "subject-visit" level, ensuring that all audio recordings from a single patient visit were exclusively assigned to either the training or the validation set within each fold. This strict separation prevents any form of data leakage and provides a more realistic estimate of the model's performance on unseen subjects.

**Optimal Threshold Determination.** Rather than employing a fixed classification threshold of 0.5, we determined an optimal threshold for each fold independently. This was achieved by constructing a Receiver Operating Characteristic (ROC) curve on the validation set and identifying the threshold that maximized Youden's J statistic (Sensitivity + Specificity - 1). This data-driven approach ensures that the model achieves an optimal balance between sensitivity and specificity for each fold, enhancing its potential clinical utility.

**Performance Metrics and Statistical Analysis.** Model performance was evaluated using a comprehensive suite of metrics, including the Area Under the ROC Curve (AUC), sensitivity, specificity, and accuracy. To quantify the statistical uncertainty of our performance estimates, we calculated 95% CIs for these key metrics by applying the non-parametric bootstrap method with 1000 resamples to the pooled predictions from all folds.

**Algorithm 1:** Model Training and Evaluation Protocol

**Input:** Full dataset $\mathcal{D}$ of subject-visits, number of folds $K = 5$, number of epochs $E = 100$, augmentation factor $A$.
**Output:** Aggregated performance metrics (AUC, Sensitivity, Specificity) with 95% CIs.

```
AllProbs ← [ ], AllLabels ← [ ]
for k = 1 to K do
    D_train, D_val ← Split D into stratified train/validation sets based on "subject-visit".
    Dataset_train ← Create dataset from D_train with data augmentation (factor A).
    Dataset_val ← Create dataset from D_val without augmentation.
    Loader_train, Loader_val ← Create DataLoaders for both datasets.
    Initialize model M_k with Xavier uniform weights.
    Initialize AdamW optimizer O_k and CosineAnnealingWarmRestarts scheduler S_k.
    Initialize EarlyStopping mechanism.
    for e = 1 to E do
        M_k.train()
        for batch in Loader_train do
            Compute logits and Focal Loss.
            Perform backpropagation and update weights with O_k.
        end
        M_k.eval()
        Compute validation loss on Loader_val.
        Update scheduler S_k.
        if EarlyStopping condition is met then
            break
        end
    end
    M_k.eval()
    Compute probabilities P_val for all samples in Loader_val.
    τ_k ← arg max_τ(Sensitivity(τ) + Specificity(τ) − 1) using P_val.
    Append P_val to AllProbs.
    Append true labels from D_val to AllLabels.
end
Compute final AUC, Sensitivity, Specificity, etc., using AllProbs and AllLabels.
Compute 95% CIs for all metrics using the bootstrap method (1000 resamples).
```

### 2 Health Economic Model Implementation

#### 2.1 Model Structure

A state-transition Markov model was developed to evaluate the potential cost-effectiveness of the AECOPD detection system from a healthcare system perspective. The model simulates a cohort of COPD patients transitioning between four mutually exclusive health states: *Stable COPD*, *Mild AE*, *Moderate-to-Severe AE*, and *Death*. The simulation was conducted over a one-year time horizon with monthly cycles to capture the dynamic nature of the disease.

The analysis compared two strategies: (1) **Standard of Care (SoC)**, representing the current management pathway without the aid of the detection system, and (2) **System-Assisted Management**, where the detection system is used to identify early signs of exacerbations, prompting timely intervention.

#### 2.2 Model Parameters and Data Sources

The model was populated with parameters derived from published literature, clinical expert consensus, and real-world data from our center. For the Probabilistic Sensitivity Analysis (PSA), each parameter was assigned a specific probability distribution to account for uncertainty. A detailed summary of these parameters, their baseline values, distributions, and sources is provided in Table 2.

A notable feature of our model is the use of an empirical distribution for the cost of moderate-to-severe

Table 1: Hyperparameters for Optimization.

| Parameter Category | Hyperparameter | Value |
|---|---|---|
| **Optimizer** | Type | AdamW |
| | Learning Rate | 1e-4 (0.0001) |
| | Weight Decay | 0.05 |
| **Learning Rate Scheduler** | Scheduler | CosineAnnealingWarmRestarts |
| | Restart Period ($T_0$) | 5 epochs |
| | Minimum Learning Rate | 1e-6 |
| **Loss Function** | Type | Focal Loss |
| | Gamma ($\gamma$) | 2.0 |
| | Alpha ($\alpha$) | 1.0 |
| **Training Process** | Epochs | 100 (with Early Stopping) |
| | Batch Size | 16 |
| | Early Stopping Patience | 50 epochs |
| | Gradient Clipping | Max norm of 1.0 |
| **Regularization** | Dropout Rate | 0.5 |

exacerbations. This distribution was constructed from 2,341 real-world hospitalization cost data points from our center. This approach enhances the model's fidelity to local clinical and economic realities compared to relying solely on published summary statistics.

Table 2: Parameters Used in the Health Economic Model.

| Parameter Category | Parameter Description | Distribution and Value | Source |
|---|---|---|---|
| **Epidemiology** | Total COPD Population (China) | Normal($\mu$ = 99.9M, 95% CI: 76.3–135.7M) | Wang et al. (2018) [6] |
| | Monthly Exacerbation Rate | Gamma($\mu = 0.101$, range 0.051–0.205) | Calzetta et al. [3] |
| | Proportion of Severe AE | Beta($\mu = 0.168$, 95% CI: 0.142–0.206) | Calverley et al. [1] |
| | Monthly Mortality (Stable) | Beta($\mu = 0.011$, 95% CI: 0.009–0.013) | Wang et al. [5] |
| | Case Fatality (Severe AE) | Beta($\mu = 0.034$, 95% CI: 0.025–0.045) | Wang et al. [5] |
| **System Effectiveness** | Sensitivity | Beta($\mu = 0.899$, 95% CI: 0.849–0.945) | Our diagnostic model results |
| | Specificity | Beta($\mu = 0.938$, 95% CI: 0.888–0.981) | Our diagnostic model results |
| | Early Intervention Reduction | Beta($\mu = 0.20$, 95% CI: 0.10–0.30) | Expert Consensus (Assumption) |
| | Patient Adherence Rate | Beta($\mu = 0.87$, 95% CI: 0.83–0.91) | Marzolini et al. [4] |
| **Economic Costs (CNY)** | Outpatient AE Treatment | Lognormal($\mu = 319$, $\sigma = 567$) | Zhu et al. [2] |
| | Inhospital AE Treatment | Empirical Distribution($\mu = 24,373$) | Our center's real-world data |
| | System Cost (per user/year) | Fixed(150) | Assumption |
| | False Positive Event Cost | Lognormal($\mu = 100$, $\sigma = 30$) | Assumption |

#### 2.3 Model Simulation and Analysis

To comprehensively assess the impact of parameter uncertainty, a Probabilistic Sensitivity Analysis (PSA) was conducted. We performed one million Monte Carlo simulations. In each simulation, a complete set of parameters was randomly drawn from their respective distributions as defined in Table 2. The Markov model was then run for both the SoC and system-assisted scenarios to calculate outcomes for that specific parameter set. The primary outputs of the model included the net cost savings, the number of avoided hospitalizations (moderate-to-severe AEs), the number of avoided deaths, and the return on investment (ROI).

Furthermore, a one-way sensitivity analysis was performed to identify the parameters with the most significant influence on the model's outcomes. The results of this analysis are presented as a tornado diagram, which visually ranks the parameters by the magnitude of their impact on the net savings. This allows for a clear identification of the key drivers of the system's economic value.

### 3 User Questionnaire

To gather data on patient lifestyle, treatment adherence, and self-perceived symptoms, we administered a custom-developed questionnaire to the study participants. The original Chinese version of this questionnaire, which was completed by all participants, is included here for reference. An English translation is also provided.

It is important to note that the self-reported outcomes from this questionnaire demonstrated a notable variance when compared against objective clinical assessments. This discrepancy was particularly pronounced among patients with cognitive impairments, suggesting that self-reported data from this cohort should be interpreted with a degree of caution. The translation of the original questionnaire is attached below:

### COPD patient management questionnaire

#### I: Basic information

**Name:** ______________

**Age:** ______________

**Contact Information:** ___________________________

**Gender:** ☐ Male ☐ Female

#### II: Living habits

**1. Smmoking status: (Please tick)**

☐ Still smoke ☐ Used to smoke, quit now ☐ Never smoke

**2. Please fill in your smoking years and the number of packs you smoked per day::**

Smoking history: ________year Average numbers per day: ________pack

**3. Daily exercises (Please tick, multiple selections are allowed) :**

☐ No ☐ Walking ☐ Taiji ☐ Indoor exercise ☐ Breathing exercise

☐ others_________

**4. Number of exercises per week:**

☐ 1 ☐ 2-3 ☐ 4-6 ☐ Every day

**5. What foods do you have every day? (Multiple selections are allowed)**

☐ Meat/fish/egg_______x50g ☐ Rice/noodles/porridge_______ x50g

☐ Vegetables_______ x50g ☐ Fruits_______ x50g ☐ Milk/soymilk _______cups

**6. Vaccination:**

Pneumonia: ☐ Yes ☐ No ☐ Don’t remember

Flu: ☐ Every year ☐ Occasionally ☐ Never ☐ Don’t remember

#### III: Management

**1. In the past year, have you been hospitalized or received intravenous drips due to cough, sputum, shortness of breath, etc.?**

☐ Yes (_______times) ☐ No

**2. In the past month, have you felt that your shortness of breath, cough, sputum and other symptoms have worsened?**

☐ Yes ☐ No

3. **Types of drugs currently used (Multiple selections are allowed):**

1. Bronchodilator

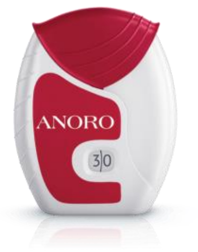


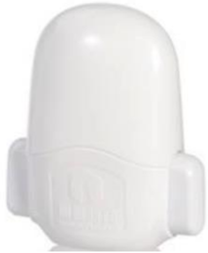

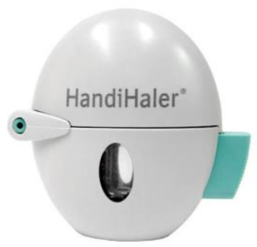


☐ ANORO ☐ Helioeast ☐ Spiriva

2. ICS

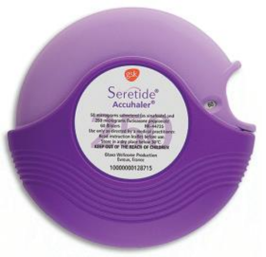


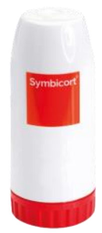


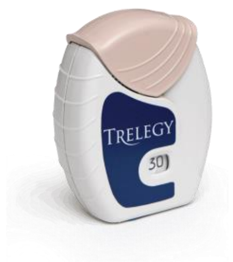


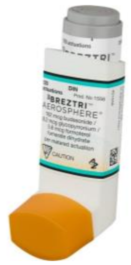

☐ Seretide ☐Symbicort ☐ Trelegy ☐ Breztri

☐ Other medications (please specify: ____________________)

**4. Frequency of inhalation:** ___ times per day __times per inhalation

**IV: Identification of AECOPD: (Please check the box that best suits your situation based on your actual experience in the past month)**

**1. Cough worsens:**

☐ Same as usual ☐ Mild ☐ Moderate ☐ Severe

**2. Increased sputum volume:**

☐ Same as usual ☐ Mild ☐ Moderate ☐ Severe

**3. Feeling that the air you breathe in is not enough:**

☐ Same as usual ☐ Mild ☐ Moderate ☐ Severe

**4. Breathing difficulties**

☐ Same as usual ☐ Mild ☐ Moderate ☐ Severe

**5. Restricted activity**

☐ Same as usual ☐ Mild ☐ Moderate ☐ Severe

### References

[1] P. M. Calverley, J. A. Anderson, B. Celli, G. T. Ferguson, C. Jenkins, P. W. Jones, J. C. Yates, and J. Vestbo. Salmeterol and fluticasone propionate and survival in chronic obstructive pulmonary disease. *New England Journal of Medicine*, 356(8):775–789, 2007.

[2] X. Chen, N. Wang, Y. Chen, T. Xiao, C. Fu, and B. Xu. Costs of chronic obstructive pulmonary disease in urban areas of china: a cross-sectional study in four cities. *International journal of chronic obstructive pulmonary disease*, pages 2625–2632, 2016.

[3] M. Miravitlles, A. D'Urzo, D. Singh, and V. Koblizek. Pharmacological strategies to reduce exacerbation risk in copd: a narrative review. *Respiratory Research*, 17:1–15, 2016.

[4] A. Paleo, C. Carretta, F. Pinto, E. Saltori, J. G. Aroca, and Á. Puelles. Mobile phone-mediated interventions to improve adherence to prescribed treatment in chronic obstructive pulmonary disease: A systematic review. *Advances in Respiratory Medicine*, 93(2):8, 2025.

[5] A. Salem, H. Zhong, M. Ramos, M. Lamotte, and H. Hu. Potential clinical and economic impact of optimised maintenance therapy on discharged patients with copd after hospitalisation for an exacerbation in china. *BMJ open*, 11(4):e043664, 2021.

[6] C. Wang, J. Xu, L. Yang, Y. Xu, X. Zhang, C. Bai, J. Kang, P. Ran, H. Shen, F. Wen, et al. Prevalence and risk factors of chronic obstructive pulmonary disease in china (the china pulmonary health [cph] study): a national cross-sectional study. *The Lancet*, 391(10131):1706–1717, 2018.
